# Extension and implementation of a system modelling the COVID-19 pandemic in Chile

**DOI:** 10.1101/2020.06.21.20136606

**Authors:** Gastón Vergara-Hermosilla, Andrés Navas

## Abstract

We modelling the dynamics of the COVID-19 epidemic taking into account the role of the unreported cases. In a first section we extend the model recently introduced/ implemented by Liu, Magal, Seydi and Webb, by considering different transmission rates for the infectious and unreported states, and we couple three new states related to hospitalized and fatalities. In addition, we introduce an operator that incorporates the effects of mitigation measures at the different rates considered in the system. Finally, we implemented the extended model in the Chilean context by considering variable the transmission rates and the fraction of unreported cases, the latter through an argument that uses mortality rates. We conclude with several conclusions and lines of future research.

## 1 Introduction

The mathematical models have played an important role in making decisions and controlling the current coronavirus epidemic. However, one of the main problems that researchers have had to face has been the lack of quality data. In particular, it is estimated that a high number of cases have been unreported, especially those of patients with low symptoms. One of the main causes of this is due to the strong demand for tests that requires a relatively complete repertorization of those infected. Indeed, the countries in which effective strategies have not been developed to increase the capacity to test the epidemic are out of control.

In general, the estimate of unreported infected has been extrapolated from the reported infected data, assuming that these numbers increase or decrease proportionally. However, it has been shown that the number of unreported cases has its own dynamics, whose role is crucial in the evolution of the epidemic [15].

In this work we address the dynamic variation of the proportions between the numbers of reported and unreported cases, incorporating them into the modelling itself.

Various mathematical models has been proposed in the literature to deal with unreported cases and their role in the progression of the epidemic. In the second section of this work, we address one of them, with the acronym SIRU, recently proposed by Zhihua Liu, Pierre Magal, Ousmane Seydi and Glen Webb [12, 13, 14]. Although the study of the qualitative theory of the underlying differential equations of the SIRU model has not been completely closed, several analogies with the classical SIR model have been concluded. However, in a previous work we have found an important difference: in the SIRU model, the case curves do not have a single peak, furthermore, a simple method has been proposed to detect parameters that originate curves with at least two peaks.

The Liu, Magal, Seydi and Webb model has already been used to describe the evolution of the epidemic in various countries (China, South Korea, the United Kingdom, Italy, France and Spain), however, in this work we extend the SIRU model, by considering different transmission rates for the infectious and unreported states, and we couple three new states related to hospitalized and fatalities cases. In addition, to obtain realistic simulations, we introduce an operator that incorporates the effects of mitigation measures at the different rates considered in the system. In the last section of this work we implement this modelling in the Chilean global scenario using the official COVID-19 data provided by the Chilean government in [16]. Unlike the implementations made in [12, 13, 14], our implementation is novel in that it uses a variable rate for disease transmission and a variable fraction of unreported cases according to the epidemic progresses, which is more relevant according to the local epidemiological reality.

The work concludes with a section dedicated to global conclusions and which describes some future lines of research. The demonstrations of the formulas referring to the basic reproductive number, the initial conditions associated with the states of infection and unreported cases, and the transmission rates associated with these states, are presented in detail in the appendices of the work.

## 2 An extension of the SIRU model

Following the approach in [15], the key assumption in this section is that unreported patients have a greater dynamic role than those that are reported (since the former do not enter into quarantine), and therefore they contribute more importantly to the epidemic. However, in the traditional SIR model, both types of patients are part of the same compartment. In [12], Liu, Magal, Seydi and Webb solve this problem by separating them into two compartments. Denoting respectively by *S, I, R* and *U* the susceptible individuals, infected individuals who do not yet have symptoms (and are at incubation stage), reported infected individuals, and unreported (either asymptomatic or low symptomatic) infected individuals, they consider the following diagram flux:

**Figure 1:**
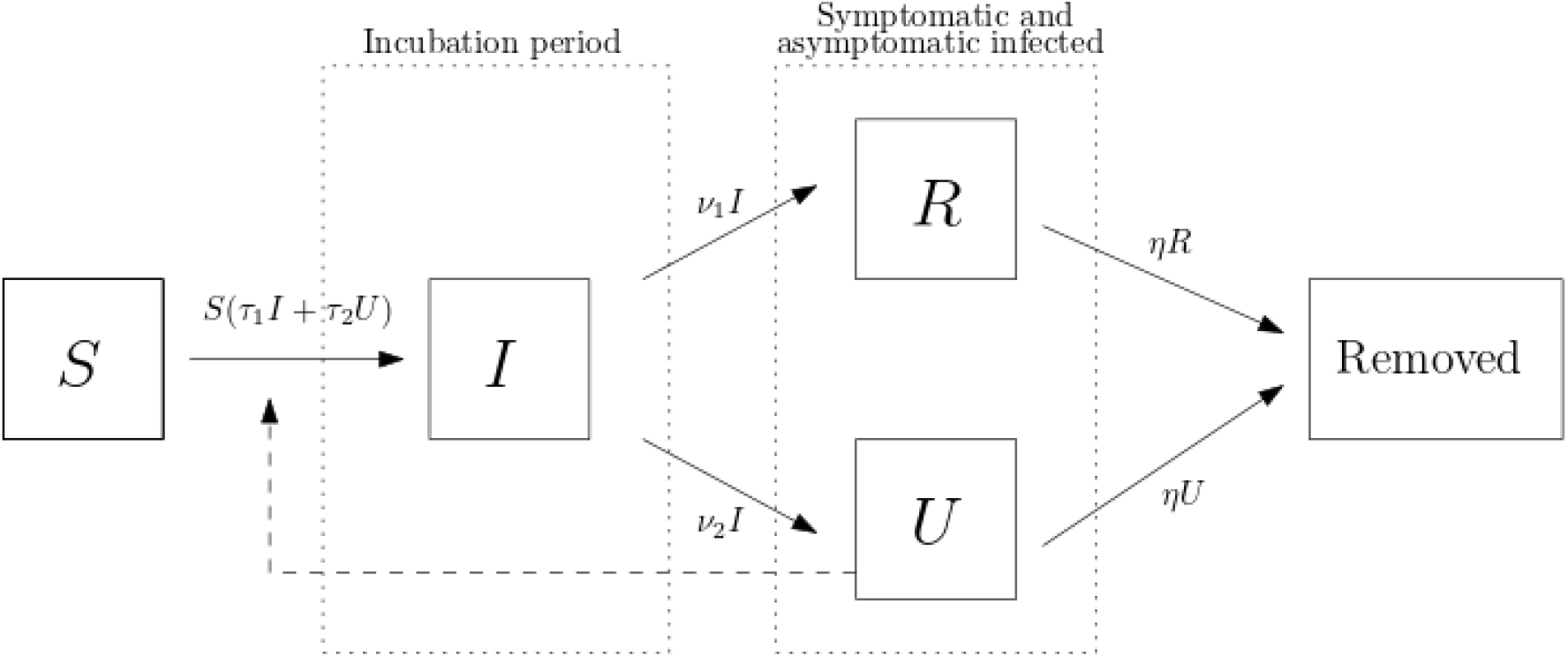
Diagram flux.

While Liu et al. in [12] assume that the parameters *τ*_1_ and *τ*_2_ related to the transmission rates of infected and unreported, respectively, are the same, in this work we do not assume this assumption.

The differential equations attached to the diagram above and that govern the dynamics of the epidemic are the following:

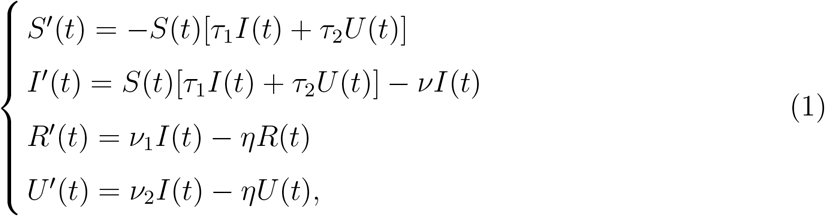

where *t ≥ t*_0_ corresponds to time, with *t*_0_ being the starting date for the study (as in [12, 13, 14], in the implementation, we will consider the time *t*_0_ corresponding to the beginning of the epidemic). Although this system of differential equations makes perfect sense when prescribing any initial condition, in epidemiological modeling one is naturally lead to use data of the following type:

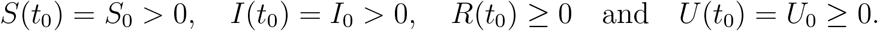

The system (1) is coupled with the following equations

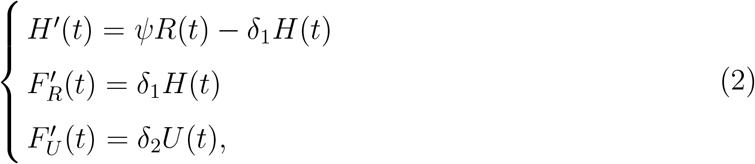

where the state *H* denote the hospitalized cases, *F*_*R*_ denote the cumulated fatalities cases reported in the official numbers and *F*_*U*_ denote the cumulated fatalities cases unreported in the official numbers.

The parameters used in the model are described in the Table below. In particular, note that *ν* = *ν*_1_ + *ν*_2_. In addition, all the parameters that are considered *τ*_1_, *τ*_2_, *ν, ν*_1_, *ν*_2_, *η, ψ, δ*_1_, *δ*_2_ are positive.

The new parameters included in our model extended are described in the following Table:

**Table 1:** Parameters and initial conditions of the model (1).

| Symbol | Interpretation |
| --- | --- |
| $t_0$ | Initial time. |
| $S_0$ | Number of individuals susceptible to the disease at time $t_0$. |
| $I_0$ | Number of infected individuals (in incubation period) without symptoms at time $t_0$. |
| $R_0$ | Number of reported infected individuals at time $t_0$. |
| $U_0$ | Number of unreported infected individuals at time $t_0$. |
| $\tau_1$ | Transmission rate of the disease. |
| $\tau_2$ | Transmission rate of the unreported infected. |
| $1/\nu$ | Average time during which the infectious asymptomatic individuals remain in incubation. |
| $f$ | Fraction of asymptomatic infected individuals that become reported infected. |
| $\nu_1 = f\nu$ | Rate at which asymptomatic infected cases become reported. |
| $\nu_2 = (1-f)\nu$ | Rate at which asymptomatic infected become unreported infected individuals (asymptomatic or mildly symptomatic). |
| $1/\eta$ | Average time during which an infected individual presents symptoms. |

Note that in the first of the equations of (1), namely

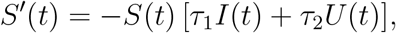

the role of *I* and *U* in the spread of the infection are differents. This is justified, for instance, by that *U* includes individuals with low symptomaticity who can practice self-care.

**Table 2:** Parameters of the extension of the model given by (2).

| Symbol | Interpretation |
| --- | --- |
| $1/\psi$ | Average time during which an infected individual with symptoms is hospitalized |
| $1/\delta_1$ | Average time during which an infected individual hospitalized dies |
| $1/\delta_2$ | Average time during which an infected individual unreported dies |

## 3 Numerical results based on official numbers

### 3.1 General implementation of the SIRU model

The *cumulative number* of infectious cases reported at a time *t*, denoted by *CR*(*t*), is given by

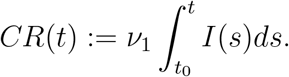

This is data that is openly available. Likewise, the cumulative number of unreported cases at a time *t* is

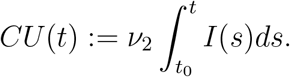

Following the scheme used by Liu, Magal, Seydi and Webb, we will assume that, at an early stage of the disease, *CD*(*t*) has an (almost) exponential form:

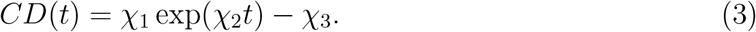

For simplicity, we will assume that *χ*_3_ = 1. The values of *χ*_1_ and *χ*_2_ will then be adjusted to the accumulated data of cases reported in the early phase of the epidemic^3^ using a classical least squares method (after passing to logarithmic coordinates). According to the above (see [12] for details), for numerical simulations, the initial time for the beginning of the exponential growth phase is fitted at

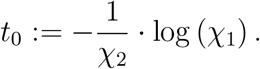

Again, for simplicity, we will identify the initial value *S*_0_ to that of the total population of Chile (since there is no prior immunity against the virus). Once the values of *ν*, and *f* are set, the conditions at the beginning of the disease are naturally fitted as

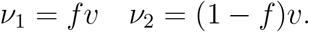

Following the approach described in the appendix, we obtain

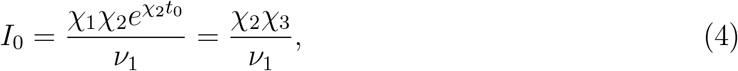

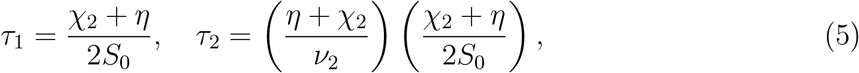

and

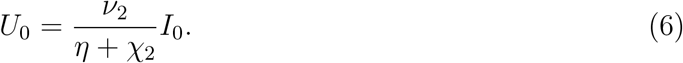

In the same form, by using the methods described in the supplementary material, the basic reproductive number for model (1) is given by

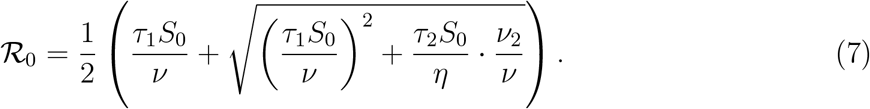

**Remark 3.1**. According to the equation (5), the parameters *τ*_1_ and *τ*_2_ are proportional, for this reason in the following to obtain estimations on the transmission rates we just consider the parameter *τ*_1_.

### 3.2 On the fraction of unreported cases

To implement the SIRU system we still need to establish a good value for the parameter *f* (the fraction of symptomatic cases that are reported). Once this is fitted, we will have the values of

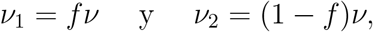

and we will just need to fit the value of *τ*.

Before continuing, it is worth pointing out that in [12, 13, 14] there is no major discussion on the criterion used to establish the value of *f* in the different scenarios. Actually, a value issued by the medical counterpart is assumed as valid. (For example, *f* = 0.8 is considered for China.) In our modeling, we will use the work of Baeza-Yates [5] and that of Castillo and Pastén [8], who use the case fatality rate of the disease (with the correct correction according to its duration; see [3, 10]) to give estimates for the right number of infected individuals.^4^ Since Baeza-Yates’ argument is simpler and is not included in an academic publication, we borrow it below in a language closer to that of the SIRU model. As we will see, it yields a method to adjusting the value of *f* that can be used in almost all contexts.

As we propose in [15], the value of *f* according to the local reality of the pandemic can be recast by

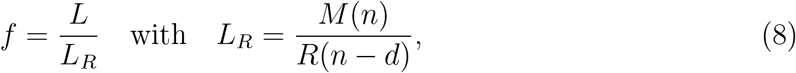

where *d* is the average time from diagnosis of infection to death, *M* (*n*) the number of deaths in day *n*, and *L* is the case fatality rate.

In the Chilean context, deaths in the period studied occurred within 9.4 days after the disease was reported. Adjusting *d* = 9 for data between 15 April to 14 May and *d* = 11 for data between 15 May to 8 June the computation of *L*_*R*_ is made feasible from the data available in [16].

Regarding the natural case fatality ratio *L* of the disease, it is deduced from international studies that, after adjusting it to the age distribution of the Chilean population, it should vary between 0.2% and 1%, with a very high tendency to be around 0.6%. In summary, this gives a parameter *f* varying between 0.1 and 0.5, with a high tendency to be close to 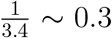. In fact, the different values obtained by using the equation (8) are shown in the following figure

**Figure 2:**
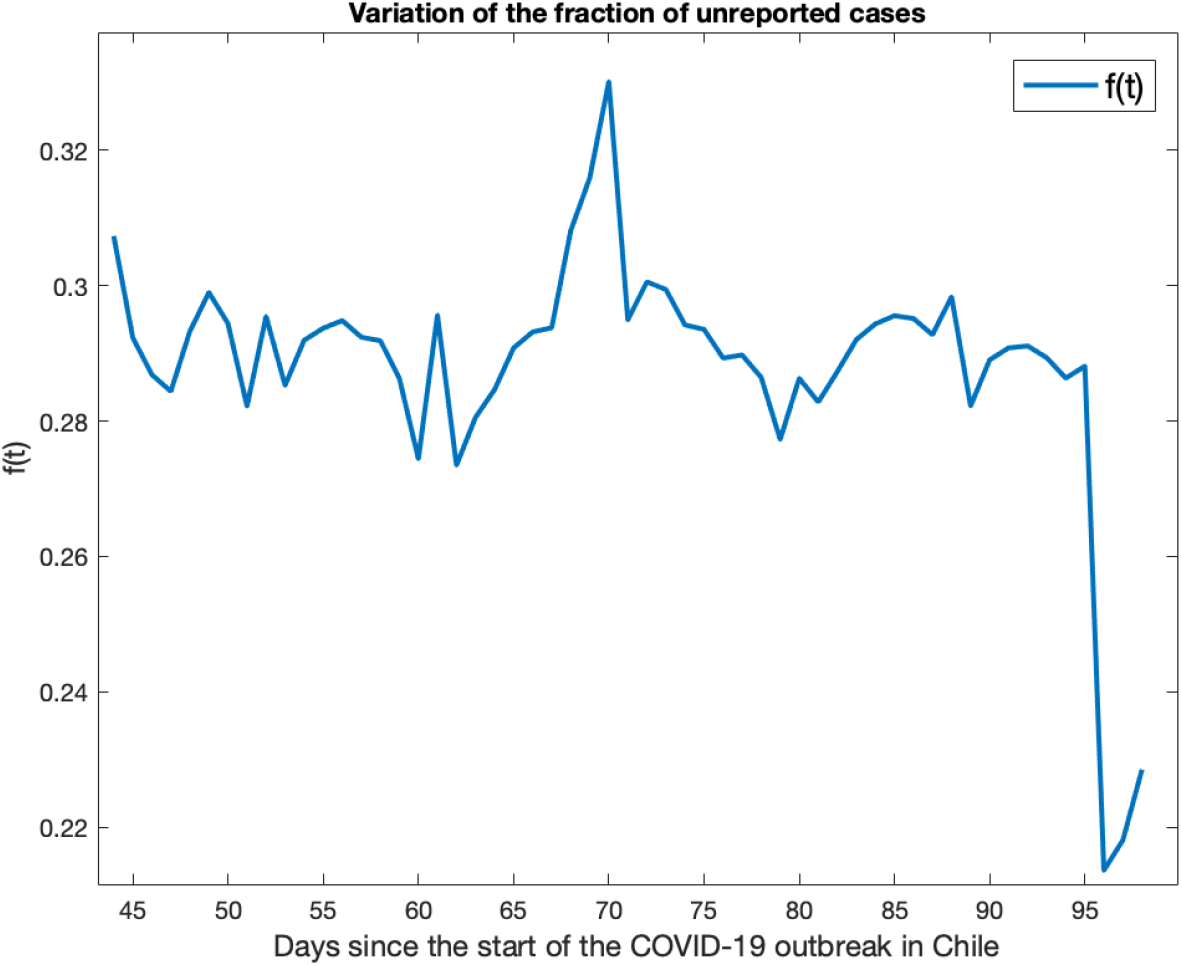
Values of *f* obtained by using the formula described in equation (8) considering *L* = 0.006.

### 3.3 Variation in the rates

Given the heterogeneity of the safeguard measures taken by the Chilean government, instead of directly applying the SIRU model, it became more pertinent to us to consider variable (in time) the different rates involucrate in the modeling process. To this end, we observe how the percentage of the Chilean population subjected to confinement has been changing, which is illustrated below:

**Figure 3:**
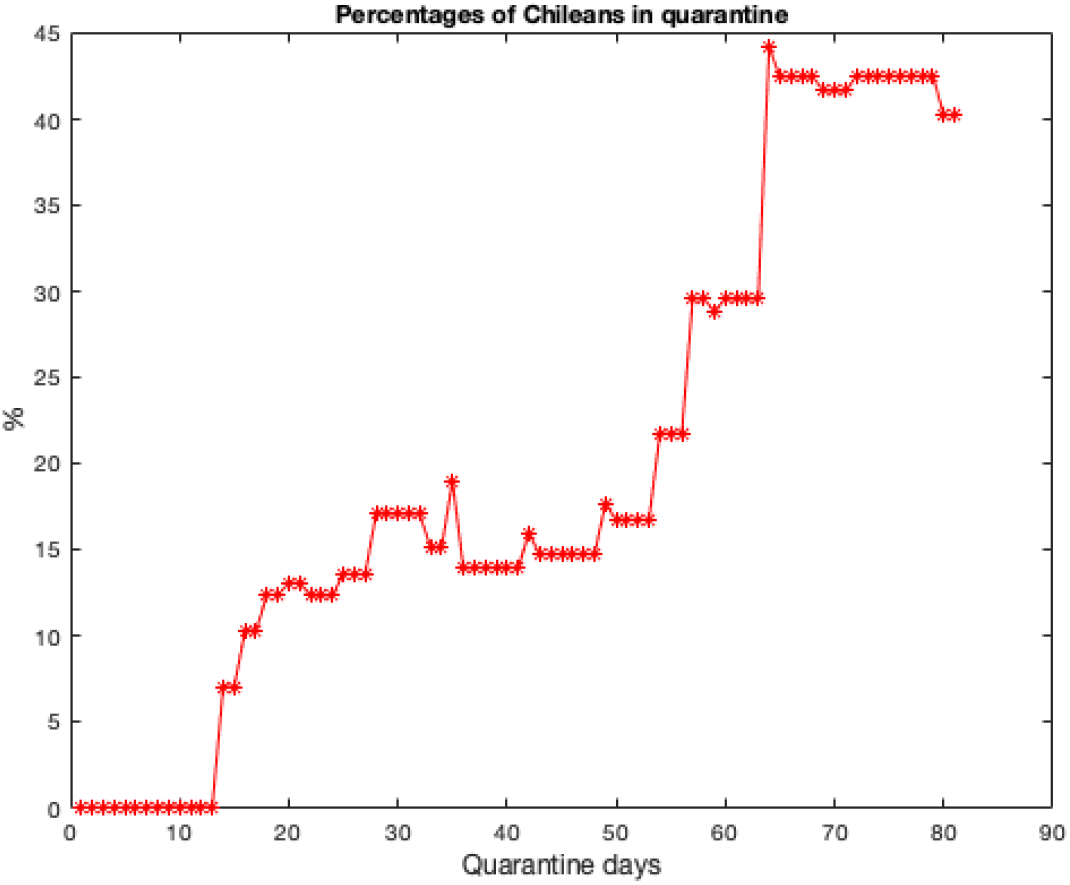
Variation of the percentages of the population in quarantine in Chile corresponding to the first 85 days from March 13, 2020.

We hence propose the following function that catches the effects of the mitigation measures taken by the government of the study area

**Definition 3.2**. The quarantine function related to the model (1) and the rate *i ∈ {τ*_1_, *ψ, δ*_1_*}*, denoted by Cuar_*i*_(*t*), is define by the following step function

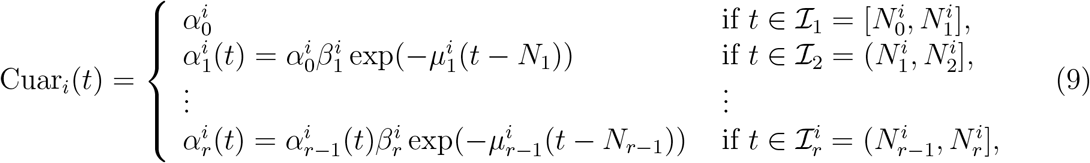

where the 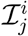’s correspond to successive time intervals and the parameters 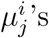 are chosen in such a way that the reported cumulated cases in the numerical simulation align with the data of the reported cumulative number of infections at time *t*.

### 3.4 Model parameters estimation and implementations

The parameter *τ*_1_ (and hence *τ*_2_) referent to the rate of transmission is estimated in the first step of the epidemy by using equation (5), however, according to Definition 3.2, the different parameters used to include the efects of the safeguard measures through the function Cuar_*τ*_1 (*t*) on the rate of hospitalized are shown in the following Table:

**Table 3:** Parameters used in Cuar_*τ*_ (*t*) to include the efects of the safeguard measures on the rate of transmission *τ*_1_.

| $j$ | $\beta_j^{\tau_1}$ | $\mu_j^{\tau_1}$ | $\mathcal{I}_j^{\tau_1}$ | $N_j^{\tau_1}$ |
| --- | --- | --- | --- | --- |
| 0 | — | — | [3 March, 22 march] | 2 March |
| 1 | 1 | $6.3 \cdot 10^{-4}$ | [23 March, 1 April] | 22 March |
| 2 | 1 | $9.1 \cdot 10^{-5}$ | [2 April, 11 April] | 1 April |
| 3 | 1 | $7.4 \cdot 10^{-5}$ | [12 April, 21 April] | 11 April |
| 4 | 1 | $6.1 \cdot 10^{-5}$ | [22 April, 1 May] | 21 April |
| 5 | 1 | $5.4 \cdot 10^{-5}$ | [2 May, 11 May] | 1 May |
| 6 | 1 | $5 \cdot 10^{-5}$ | [12 May, 21 May] | 11 May |
| 7 | 1 | $4 \cdot 10^{-5}$ | [22 may, 1 June] | 21 May |
| 8 | 1 | $1.7 \cdot 10^{-4}$ | [2 June, 9 June] | 1 June |

The parameter *ψ* referent to the rate of hospitalized is estimated as

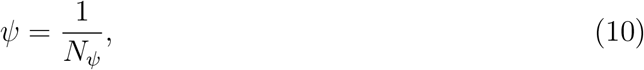

where *N*_*ψ*_ is the hospitalized rate and *p*_*ψ*_ is the case hospitalized rate (namely the fraction of hospitalized per reported infectious individuals). In the simulation we chose *N*_*ψ*_ = 3 days, however, according to Definition 3.2, the different parameters used to include the efects of the safeguard measures through the function Cuar_*ψ*_(*t*) on the rate of hospitalized *ψ* are shown in the following Table:

**Table 4:** Parameters used in Cuar_*ψ*_(*t*) to include the efects of the safeguard measures on the rate of hospitalized *ψ*.

| $j$ | $\beta_j^\psi$ | $\mu_j^\psi$ | $\mathcal{I}_j^\psi$ | $N_j^\psi$ |
| --- | --- | --- | --- | --- |
| 1 | 3 | $5.5 \cdot 10^{-2}$ | [15 April, 21 April] | — |
| 2 | 3 | $2.4 \cdot 10^{-2}$ | [22 April, 1 May] | 21 April |
| 3 | $3 \cdot 0.49$ | $1 \cdot 10^{-3}$ | [2 May, 11 May] | 1 May |
| 4 | $3 \cdot 0.49$ | $-3 \cdot 10^{-3}$ | [12 May, 21 May] | 11 May |
| 5 | 1.5 | $2.1 \cdot 10^{-2}$ | [22 may, 1 June] | 21 May |
| 6 | 1.206 | $2.5 \cdot 10^{-2}$ | [2 June, 9 June] | 1 June |

In the case of the fatalities parameters *δ*_1_ and *δ*_2_, these are estimated as

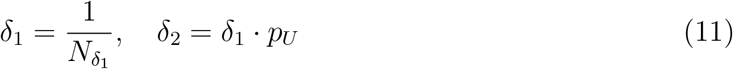

where 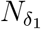 is the fatalities rate, and *p*_*U*_ is a parameter of uncertainty related to the increase of fatalities in the official civil register of the goverment of Chile not caused a priori by COVID-19 (for details see [17]). In the simulation we chose *p*_*U*_ = 1*/*10 and 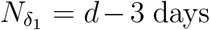, where *d* is the average time from diagnosis of infection to death. Specifically, we consider *d* = 9 for data between 15 April to 14 May and *d* = 11 for data between 15 May to 8 June.

**Table 5:**
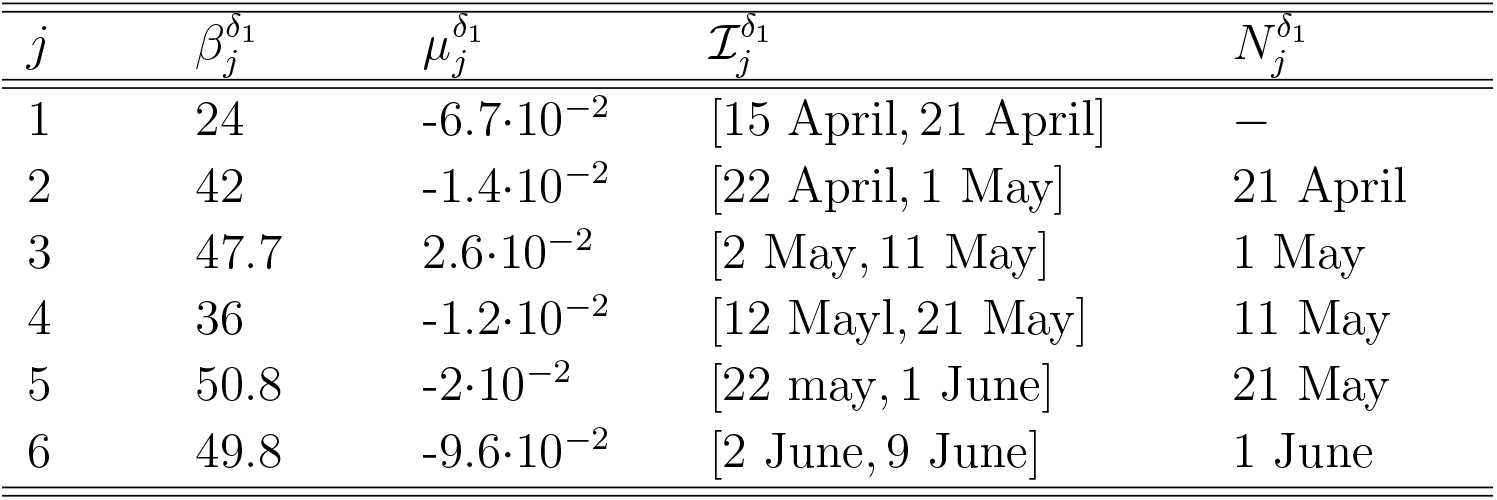
Parameters used in 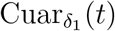 (*t*) to include the efects of the safeguard measures on the rate of transmission *δ*_1_.

Using the data of the cumulated reported cases, hospitalized and fatalities available in [16], we can finally proceed to the simulations, which are shown below. They illustrate numerical estimates for the curves of *CR*(*t*), *CU* (*t*), *H*(*t*), *F*_*R*_(*t*) and *F*_*U*_ (*t*).

**Figure 4:**
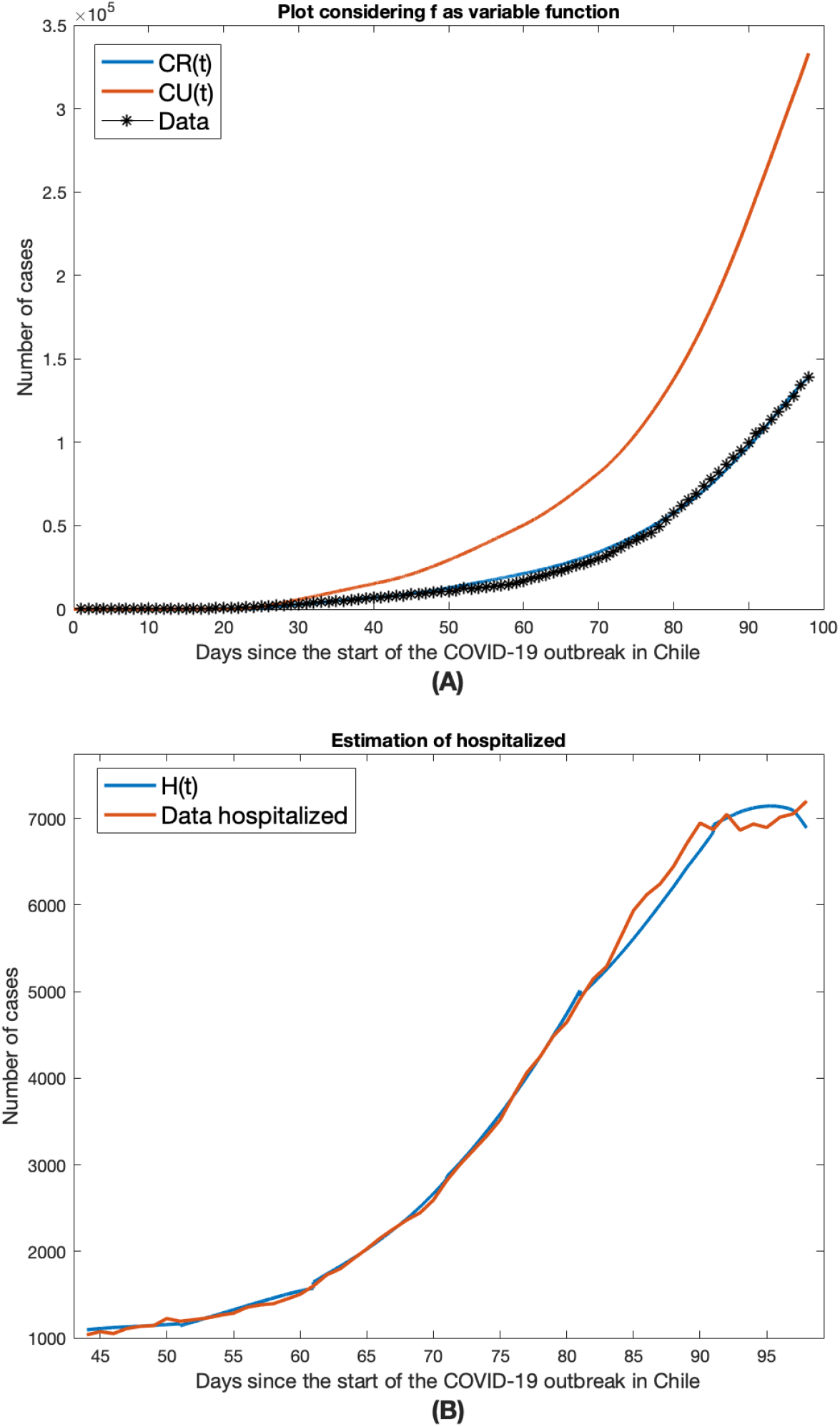
Plots of the numerical approximations of the functions *CR*(*t*), *CU* (*t*) and *H*(*t*) obtained from the numerical solutions of the model (1) coupled with (2) applied to the Chilean context based on the data of reported cumulated cases and hospitalized up to 8 June 2020 [16]. The plots (A) and (B) were obtained by considering *f* variable in time, using the parameters *η* = 1*/*7 and *ν* = 1*/*7.

**Figure 5:**
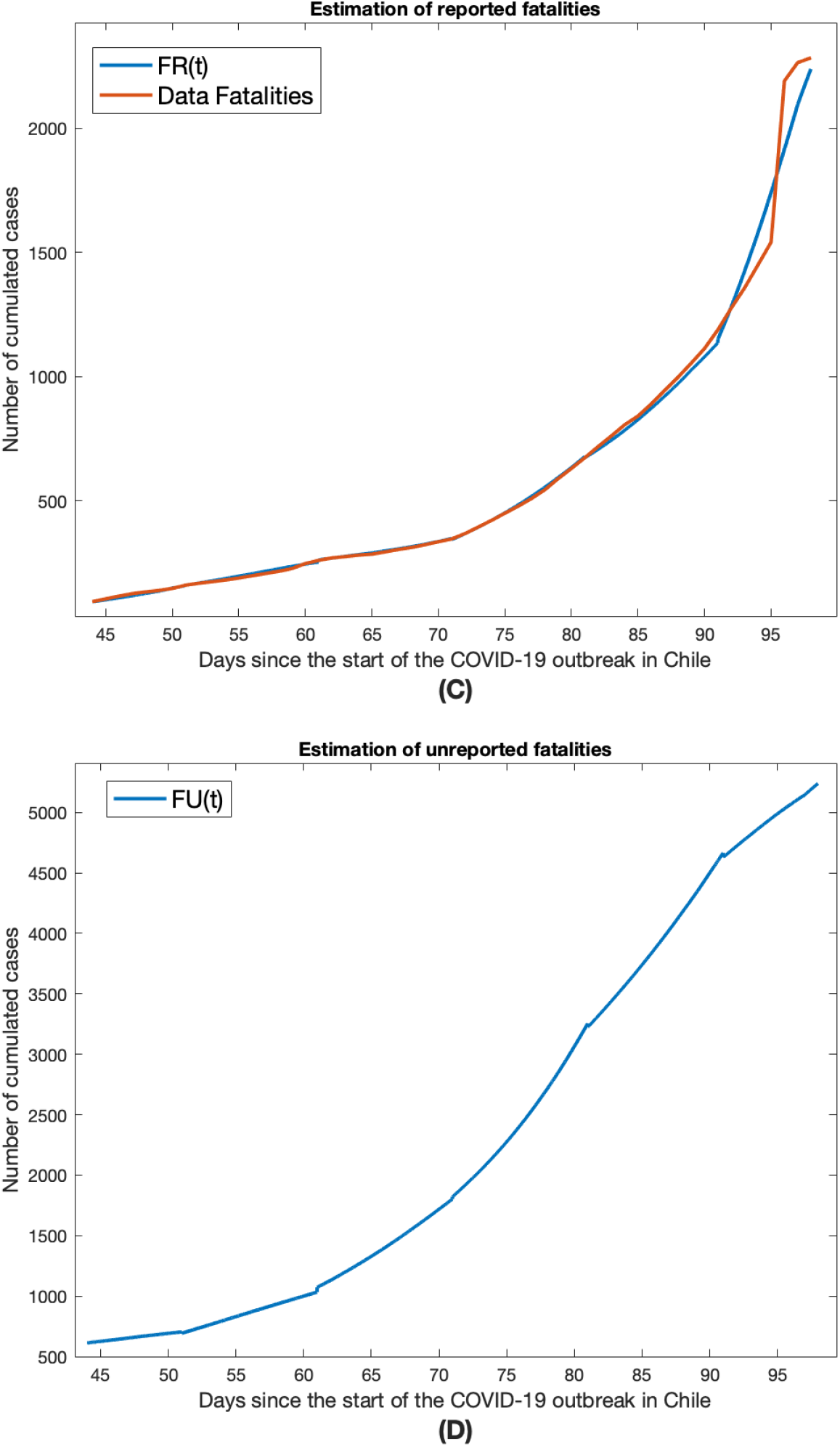
Plots of the numerical approximations of the functions *F*_*R*_(*t*) and *F*_*U*_ (*t*) obtained from the numerical solutions of the model (1) coupled with (2) applied to the Chilean context based on the data of reported cumulated cases up to 8 June 2020 [16]. The plots (C) and (D) were obtained by considering *f* variable in time, using the parameters *η* = 1*/*7 and *ν* = 1*/*7.

## 4 Discussion and future work

The epidemic outbreak of the new coronavirus COVID-19 was first detected in Wuhan, China, in late 2019. In Chile, the first case was reported on March 3, 2020, in Talca. Since then, modeling the epidemic in the country has faced the problem of the low availability of disaggregated data [4].

The first part of the work naturally led us to model the dynamics of the epidemic incorporating a compartment for unreported cases so that we could deal with their active role in the evolution. To do this, we consider the SIRU model recently introduced/implemented by Liu, Magal, Seydi and Webb in [12, 13, 14], and we extend it considering different transmission rates for the infected and unreported states and coupling three new states, referents to hospitalized and fatalities.

In the second part of the work, we implemented the extension of the SIRU model to the case of Chile. The simulations carried out considering an extension of the model of Liu et al. are based on the data of cases reported in the official numbers from March 3 to June 8, 2020. Our extension of the system that includes estimates on the number of hospitalized and dead has been considered in a shorter time interval, this is mainly due to the fact that the data regarding the number of hospitalized people began to be published by the Chilean government in [16] from April 15. Regarding the method used to carry out our implementations, we consider variable transmissions rates for infected and unreported cases, which is more appropriate according to the local reality. Our rate is coupled to the official statistics provided by the government in [16], information that also allowed us to make the parametric ajustement. An important ingredient to launch the simulations was the value of the fraction *f* which now is considered variable and is obtained following the approach introduced by Navas and Vergara-Hermosilla in the preliminary work [15]. In the model extended the estimate of fatalities are divided in two disjoint groups: reported fatalities and unreported fatalities. The main reason for consider this assumption is the fact that in the last month the Chilean health system was collapsed, and in several cases the deads are not incluided in the officials numbers on COVID-19, increasing without apparent explanation the number of deceased in the official civil registries of the government of Chile. On the other hand, we incorporate the operator Cuar_*i*_(*t*) which is such includes the effects of the different mitigation measures taken for the Chilean government on the transmission, hospitalization, and death rates, providing a more realistic simulation of the epidemic.

Although the estimates above were obtained *a posteriori*, their complementarity puts us in a good position to use them for modeling the future evolution of the epidemic. We strongly believe that the basis for pursuing the implementation of the SIRU model are fullfilled, and it would be very useful to advance in a more compartmentalized and georeferenced implementation of it.

## Data Availability

Official data of Covid-19 by the Government of Chile

### Appendix 1: materials and methods

By using equation (3), we obtain

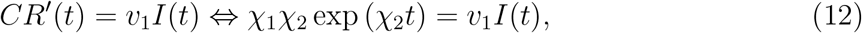

and hence, by evaluating the last equation at *t* = *t*_0_, we deduce that

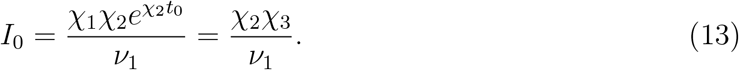

Moreover, by consider equation (3), we obtain

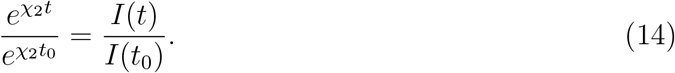

and then, we conclude that

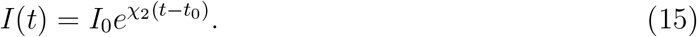

In order to evaluate the parameters of the model, we replace *S*(*t*) by the total population in the zone considered on the right-hand side of (1) (which is equivalent to neglecting the variation of susceptibles due to the epidemic, which is consistent with the fact that *t → CR*(*t*) grows exponentially). Therefore, it remains to estimate the parameters *τ*_1_ and *τ*_2_ in the following linearized system:

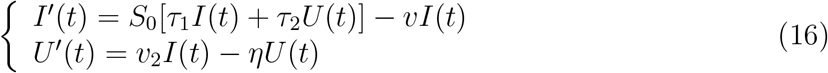

By consider the first equation, we obtain

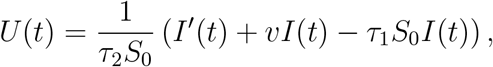

and therefore, by using (15), we must have

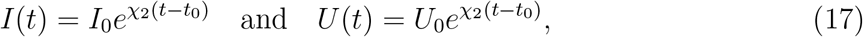

hence, by reemplacing these expressions into (16), we obtain

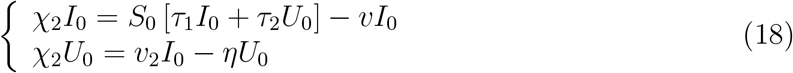

By dividing the first equation of (18) by *I*_0_ we obtain

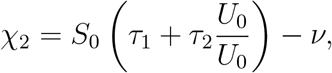

and then

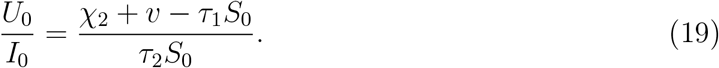

By using the second equation of (18), we obtain

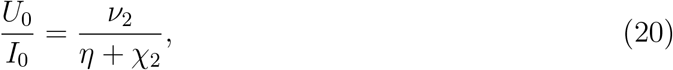

and then, we conclude that

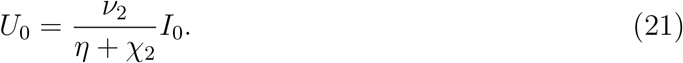

Now, by consider by linearizing the first equation of the model (1), we obtain

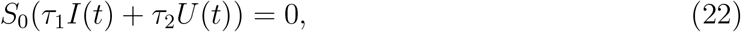

and, by using (17) and (21) on (22), we deduce

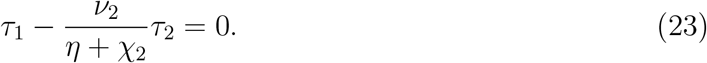

Moreover, by using (19) and (20), we obtain

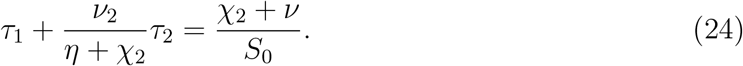

Then, by solve the equations (23) and (24) for *τ*_1_ and *τ*_2_, we deduce

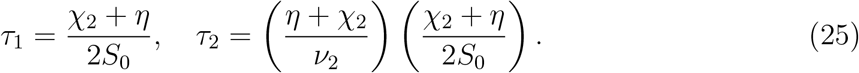

#### 4.1 Computation of basic reproductive number *R*_0_

In this section we consider classic results on the basic reproductive number (see for instance, Chapter 7 on the book of Dieckmann, Heesterbeek and Britton [2]). The linearized equation of the infectious part of the model is given by

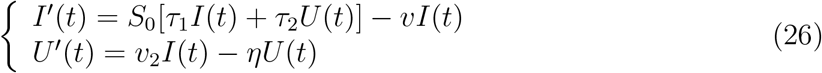

The corresponding matrix is

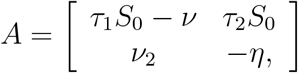

which can be rewritten as *A* = *V − S*, where

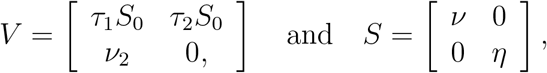

and then, the next generation matrix is given by

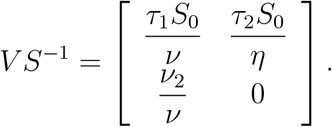

Then, by denoting by *ρ*(*M*) the spectral ratio of the matrix *M*, and by consider equation 7.18 of Chapter 7 in [2], the basic reproductive number is given by

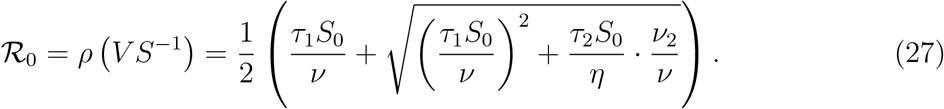

## Footnotes

3 Specifically, to adjust *χ*^1^, *χ*^2^ we consider the next 28 days since the detection of the first case in Chile (Talca).

4 They estimate this number between 60% and 70% higher than the one reported for the period studied.

## Notes

1 Supported by the European Union’s Horizon 2020 research and innovation programme under the Marie Sklodowska-Curie grant agreement No 765579.

2 Supported by MICITEC Chile.

### Competing Interest Statement

The authors have declared no competing interest.

### Funding Statement

Supported by the European Union Horizon 2020 research and innovation programme under the Marie Sklodowska-Curie grant agreement No 765579.

### Author Declarations

IMB, Universite de Bordeaux

